# Impact of imperfect diagnosis in ME/CFS association studies

**DOI:** 10.1101/2022.06.08.22276100

**Authors:** João Malato, Luís Graça, Nuno Sepúlveda

**Affiliations:** Instituto de Medicina Molecular João Lobo Antunes, Faculty of Medicine, University of Lisbon, Lisbon, Portugal; CEAUL–Centro de Estatística e Aplicações da Universidade de Lisboa, University of Lisbon, Lisbon, Portugal; Faculty of Mathematics and Information Science, Warsaw University of Technology, Warsaw, Poland

**Keywords:** misdiagnosis, misclassification, association studies, simulation, power studies, myalgic encephalomyelitis, chronic fatigue syndrome

## Abstract

The absence of an objective disease biomarker puts studies of Myalgic Encephalomyelitis/Chronic Fatigue Syndrome (ME/CFS) under the curse of imperfect diagnosis. This problem leads to frequent reports that fail to reproduce prior published studies. To address the impact of imperfect diagnosis on the robustness of studies’ conclusions, we conducted a simulation study to quantify the statistical power to detect a disease association with a hypothetical binary factor in the presence of imperfect diagnosis. Using the classical case-control design, studies with sample sizes of less than 500 individuals per group could not reach the target power of at least 80% to detect realistic disease associations. We then recreated serological association studies in which the chance of imperfect diagnosis was combined with the probability of misclassifying a binary factor, as it happens in a typical serological association study. In this case, the target power of 80% could only be achieved for studies with more than 1000 individuals per group. Given the current sample sizes of ME/CFS studies, our results suggest that most studies are likely to be underpowered due to imperfect diagnosis alone. To increase reproducibility across studies, we provided some practical recommendations, such as the use of standard case definitions together with multi-centric study designs, and routine reporting of power calculations under a non-negligible chance of misdiagnosis. Our results can also inform the design of future studies under the assumption of misdiagnosis.

## 1 INTRODUCTION

Myalgic encephalomyelitis/chronic fatigue syndrome (ME/CFS) is a complex disease whose patients manifest unexplained long-lasting fatigue or post-exertional malaise upon minimal physical or mental effort (1, 2). The disease has a sex bias towards women, with a ratio of about two women to one man among cases (3). The real burden of the disease remains unknown, but current prevalence estimates range from 0.1% to 2.2% (4).

Given the lack of a biomarker for the disease, the diagnosis of suspected cases is made by a set of key symptoms together with the exclusion of known diseases, such as multiple sclerosis and diabetes, which could explain fatigue (5). To make the disease diagnosis even more cumbersome, there are multiple case definitions proposed in the literature (6). A possible solution to this problem is to follow expert consensus recommendations for clinical and research practices. In this regard, the European Network on ME/CFS (EUROMENE) suggests the use of the Centre for Disease Control (Fukuda/CDC) criterion (1) and the Canadian Consensus Criteria (CCC) (2) for research purposes (7). In a clinical setting, the ME/CFS diagnosis can also contemplate the Institute of Medicine (IOM) criterion due to its simplicity (8, 9).

The use of different case definitions by clinicians and researchers opens the door to not identifying the same population of patients. As a consequence, it is difficult to ascertain the epidemiology and the real economic costs of the disease (4, 7). Such a discrepancy in case identification has been demonstrated in a study where less than 75% of patients had the same diagnostic outcome across Fukuda/CDC, CCC, and IOM criterion (10). In the same study, data from a symptom assessment questionnaire alone could not completely separate patients diagnosed with ME/CFS from healthy controls and patients with multiple sclerosis. Similar symptoms overlap was found between ME/CFS, multiple sclerosis (11, 12), and rheumatoid arthritis (13); the exclusion of these two autoimmune diseases is, however, part of current ME/CFS diagnostic criteria. In this scenario, current ME/CFS diagnostic criteria are likely to have different sensitivities and are far from 100%.

In general, imperfect diagnostic tests imply the presence of estimation bias in subsequent statistical inference. It also reduces the statistical power to detect true disease associations or the efficiency of estimating the true prevalence (14, 15). These implications are possible explanations for the lack of scientific reproducibility in ME/CFS research. In this regard, Nacul et al. (16) suggested missing important disease associations due to the problem of imperfect ME/CFS diagnosis (i.e., misdiagnosis). However, this study did not include a systematic analysis of the potential impact of misdiagnosis on ME/CFS-related associations. In this scenario, we aimed to investigate the statistical power of hypothetical case-control association studies of ME/CFS in the presence of misdiagnosis. For this purpose, we performed an extensive simulation study under different sample sizes, levels of misdiagnosis, and strength of disease associations. We also investigated the additional impact of sensitivity and specificity associated with serological evaluations that are often performed in ME/CFS (17, 18, 19). Finally, we extended our study to discuss data from two publications (20, 21).

## 2 STATISTICAL METHODOLOGY

### 2.1 Statistical formulation of the problem

To study the impact of misclassification, let us assume a classical case-control association study in which diagnosed ME/CFS patients and healthy controls were matched for possible confounders such as age, gender, or body mass index (BMI). The main objective of this study is to investigate a potential association between a candidate causal factor (e.g., a genetic factor or the occurrence of a given infection) and ME/CFS. For simplicity, let us assume that this factor has only two possible outcomes: present versus absent. This is the classical epidemiological situation of a putative risk factor in which individuals can be divided into exposed and non-exposed. The hypothetical data can be summarised by a 2 × 2 contingency table, whose sampling distribution is the product of two independent Binomial distributions, one Binomial distribution per group,

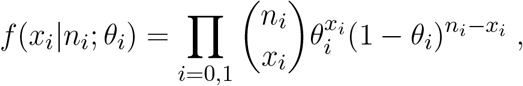

where *x*_0_ and *x*_1_ are the observed frequencies of healthy controls and suspected cases with presence of the candidate causal factor, respectively, *n*_0_ and *n*_1_ are the corresponding sample sizes of each group, and *θ*_0_ and *θ*_1_ are the probabilities for the presence of the candidate causal factor in healthy controls and suspected cases, respectively.

To investigate whether there is evidence for an association between this candidate factor, one can perform a hypothesis test in which the null and alternative hypotheses, *H*_0_ and *H*_1_, are

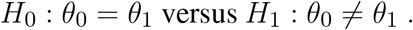

The same hypotheses can be rewritten in terms of the odds ratio as follows

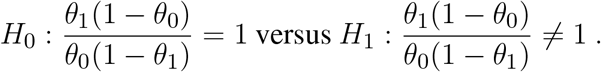

To test these hypotheses, one can use the Pearson’s χ^2^ test for independence in 2-way contingency tables. Using a significance level of 0.05, p-values < 0.05 suggest a rejection of the null hypothesis.

To study the impact of misdiagnosis on the power of above Pearson’s χ^2^ test, we considered seven simplifying assumptions for the statistical formulation of the problem:

I. Suspected cases are composed of both apparent and true patients;
II. The presence of the candidate causal factor is only associated with true patients;
III. Apparent cases are similar to healthy controls in what the association with the candidate causal factor is concerned;
IV. The chance of making a ME/CFS misdiagnosis is only dependent on the true clinical status of the cases and not on the confounders;
V. The association to be detected is independent of disease duration and disease triggers, among other factors occurring during the disease course;
VI. There is no misdiagnosis (or misclassification) of the true health status of the controls;
VII. The presence or absence of the candidate causal factor is determined accurately in each individual.

Assumption VI is invoked to exclude the (unlikely) situation in which healthy controls could include true undiagnosed ME/CFS patients. Assumption VII is appropriate for genetic association studies in which the genotypic error rate is quantified and typically low (22).

The above assumptions allow, then, to rewrite the probability of the causal factor being present in suspected cases as

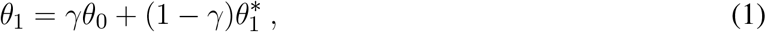

where *γ* is the misdiagnosis (or misclassification) probability, and 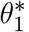 is the probability of the candidate causal factor being present in true ME/CFS cases. The direct consequence of the above formula is a dilution effect of the association operated by the possibility of misdiagnosis. If misdiagnosis could be directly observed, the classical 2 × 2 contingency table could be augmented as in Table 1, where the diagnosed cases are divided into apparent and true cases in accordance with assumption I.

**Table 1.** Augmented version of the observed 2 × 2 contingency table in presence of misdiagnosis of ME/CFS cases for a classical case-control association study.

| Causal factor | Controls | ME/CFS-diagnosed cases |  |
| --- | --- | --- | --- |
|  |  | (Apparent) | (True) |
| Present | $\theta_0$ | $\gamma\theta_0$ | $(1 - \gamma)\theta_1^*$ |
| Absent | $1 - \theta_0$ | $\gamma(1 - \theta_0)$ | $(1 - \gamma)(1 - \theta_1^*)$ |

A more-complex statistical situation is to consider the chance of misdiagnosis together with the impossibility of determining the presence of the candidate causal factor accurately. This situation is the curse of all serological studies on ME/CFS in which the objective is to evaluate the association between the disease and the presence of antibodies against a given infectious agent (e.g., Epstein-Barr virus (23)). In these studies, individuals are deemed seropositive or seronegative based on a threshold in the antibody distribution. When the estimated antibody distributions for these two populations overlap, the respective serological classification is subject to uncertainty ruled by the underlying sensitivity and specificity of the statistical method used (24). The description below follows as if the data at hand were generated from this type of study.

To model this more complex situation, previous assumption VII is replaced with the two additional assumptions:

VII. There are only two possible serological outcomes for each individual: seronegative or seropositive;
VIII. The sensitivity and specificity of the serological classification are shared across all the individuals.

The revised assumption VII aims to exclude the situation where the serological classification can contemplate an indeterminate status due to the laboratory protocol (21) or the presence of multiple serological populations (25). Similarly to assumption V for misdiagnosis, the new assumption VIII intends to disregard the effect of confounders (i.e., age or gender) and disease-related factors (i.e., disease duration or disease severity) on the performance of the serological classification.

Under the assumptions I–VIII the probability of recording the presence of the candidate causal factor in a ME/CFS patient can be generalised to

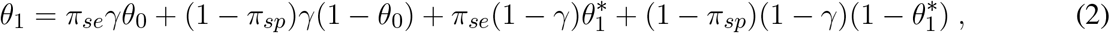

where *π*_*se*_ and *π*_*sp*_ are sensitivity and specificity for the serological classification, respectively. Note that, when *π*_*se*_ = *π*_*sp*_ = 1 (perfect serological testing), the above formula converts to equation (1). Table 1 can thus be further augmented by including the true serological status of the individuals (Table 2).

**Table 2.** Augmented version of the observable 2 ×2 frequency Table in the case-control association study with possible misdiagnosis of ME/CFS cases and misclassification of the true serological status (seropositive, *S*^+^, and seronegative, *S*^−^).

| Estimated Serological status | True serological status | Controls | ME/CFS-diagnosed cases |  |
| --- | --- | --- | --- | --- |
|  |  |  | (Apparent) | (True) |
| $S^+$ | $S^+$ | $\pi_{se}\theta_0$ | $\pi_{se}\gamma\theta_0$ | $\pi_{se}(1 - \gamma)\theta_1^*$ |
| | $S^-$ | $(1 - \pi_{sp})(1 - \theta_0)$ | $(1 - \pi_{sp})\gamma(1 - \theta_0)$ | $(1 - \pi_{sp})(1 - \gamma)(1 - \theta_1^*)$ |
| $S^-$ | $S^+$ | $(1 - \pi_{se})\theta_0$ | $(1 - \pi_{se})\gamma\theta_0$ | $(1 - \pi_{se})(1 - \gamma)\theta_1^*$ |
| | $S^-$ | $\pi_{sp}(1 - \theta_0)$ | $\pi_{sp}\gamma(1 - \theta_0)$ | $\pi_{sp}(1 - \gamma)(1 - \theta_1^*)$ |

### 2.2 Simulation study

To investigate the impact of both misdiagnosis scenarios on detecting an association between a candidate causal factor and ME/CFS, we performed a comprehensive simulation study assuming matched sample sizes for ME/CFS patients and healthy controls (*n*_0_ = *n*_1_). We considered the following sample sizes per study group: 100, 250, 500, 1000, 2500, and 5000. This choice of sample size is probably optimistic for ME/CFS research, given the sample sizes (mean = 86.2, range = 16–300 for ME/CFS group) reported in immune marker studies conducted in Europe (26).

To parameterise the simulation study, we first specified the association between the candidate causal factor and true ME/CFS patients by the odds ratio (hereafter denoted as Δ_*T*_) and the probability of the presence of the causal candidate factor in healthy controls (*θ*_0_). We considered Δ_*T*_ ∈ {1.25, 1.5, 2, 3, 5, 10}, whose values ranged from weak to strong associations. Given the lack of disease-specific biomarkers, it is expected that the putative associations to be detected should not be too strong. However, the simulation of strong associations is important to assess what to expect under an optimistic scenario. In addition, we specified *θ*_0_ ∈ {0.05, 0.1, 0.25, 0.5 }. In the case of candidate gene scenarios, *θ*_0_ could represent the minor allele frequency of a given single nucleotide variant in the healthy population. Combining the values of *θ*_0_ together with the ones specified for Δ_*T*_, we could study the impact of misdiagnosis on detecting whether ME/CFS follows the “common disease, rare variant hypothesis” or the “common disease, common variant hypothesis”. For these two contrasting hypotheses, a common disease could be the result of either multiple rare genetic factors with a high penetrance or many common genetic variants with low penetrance (27, 28). Having *θ*_0_ and Δ_*T*_ fixed in the respective values, we can determine the value of 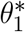 as follows

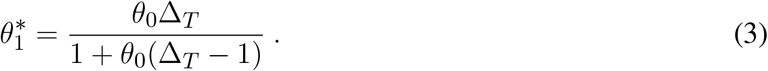

With respect to the probability of misdiagnosis, *γ*, we allowed this parameter to vary from 0 (all cases were correctly diagnosed) to 1 (all cases were incorrectly diagnosed) with a lag of 0.01.

To simulate data from the second misdiagnosis scenario, we considered fixed parameters Δ_*T*_ = 3 and *θ*_0_ = 0.25. For parameters *π*_*se*_ and *π*_*sp*_, we considered all possible combinations of 0.80, 0.90, 0.925, 0.975, and 1.0, where *π*_*se*_ = *π*_*sp*_ = 1 corresponded to the first scenario.

For each misdiagnosis scenario, parameter set, and sample size, we simulated 10,000 datasets to estimate the power of rejecting *H*_0_ when the possibility of misdiagnosis was ignored. A detailed description of the simulation procedure can be found elsewhere (10). In each data set, we rejected *H*_0_ if the p-value of the Pearson’s χ^2^ test was less than the usual level of significance, *α* = 0.05. The power of the study was estimated as the overall proportion of simulated data sets in which *H*_0_ was rejected. To better understand the practical implications of the simulation results, we specified a target power of at least 80% for the probability of rejecting the null hypothesis in favour of the alternative hypothesis, given that the latter was true. The term used for this target power is generally denoted as 1 − *β*, where *β* represents the probability of incorrectly failing to reject *H*_0_ (Type II error). A target value of 80% suggests that a true association is very likely to be reproducible in a follow-up study.

### 2.3 Application to two ME/CFS studies

We also studied the impact of misdiagnosis on available data from a candidate gene association study and an immunological evaluation study. The first study recruited 201 self-reported healthy controls and 305 ME/CFS patients whose symptoms complied with CCC (20). All participants were genotyped for 5 SNPs previously associated with autoimmune diseases:

- rs2476601, tyrosine phosphatase non-receptor type 22 (*PTPN22*);
- rs3087243, cytotoxic T-lymphocyte-associated protein 4 (*CTLA4*);
- rs1800629 and rs1799724, tumor necrosis factor (*TNF*);
- rs3807306, interferon regulatory factor 5 (*IRF5*).

The study found significant associations of rs2476601 (*PTPN22*) and rs3087243 (*CTLA4*) with ME/CFS patients potentially triggered by an acute infection.

The second study encompassed data of 251 patients and 107 healthy controls from the UK ME/CFS biobank (21). Patients complied with either Fukuda/CDC or CCC. The original objective of this study was to perform a detailed comparison between these two groups in terms of immune-cell data. Here, we focused on data of antibody positivity related to six different herpesviruses: human cytomegalovirus (CMV), Epstein-Barr virus (EBV), herpes simplex virus 1 and 2 (HSV1 and HSV2), varicella-zoster virus (VZV), and human herpesvirus (HHV6). Note that, for each virus, antibody positivity was determined by a cut-off value as recommended by the manufacturers of the serological kits. However, another re-analysis of the same data provided evidence for an overlap between the antibody distribution of hypothetical antibody-negative and antibody-positive populations (24). Therefore, besides the potential ME/CFS misdiagnosis, the subsequent antibody classification could also be affected by misclassification.

In both studies, we estimated the power of detecting an association as a function of misdiagnosis probability, *γ*, using simulated data generated from the reported associations, as explained later.

## 3 RESULTS

### 3.1 Simulation study: impact of ME/CFS misdiagnosis

The power to detect an association with ME/CFS decreased with the misdiagnosis probability (Figure 1 and Figure 2). The maximum power was achieved when all the patients were assumed to be true cases (*γ* = 0), that is, when there was no dilution effect of the true association to be detected. In the other extreme, when all the patients were assumed to be apparent cases (*γ* = 1), the corresponding power was equal to the significance level set up for the analysis (*α* = 0.05). This result was a direct consequence of assumption III, in which the misdiagnosed cases were deemed identical to healthy controls in what the association with the candidate causal factor was concerned.

**Figure 1.**
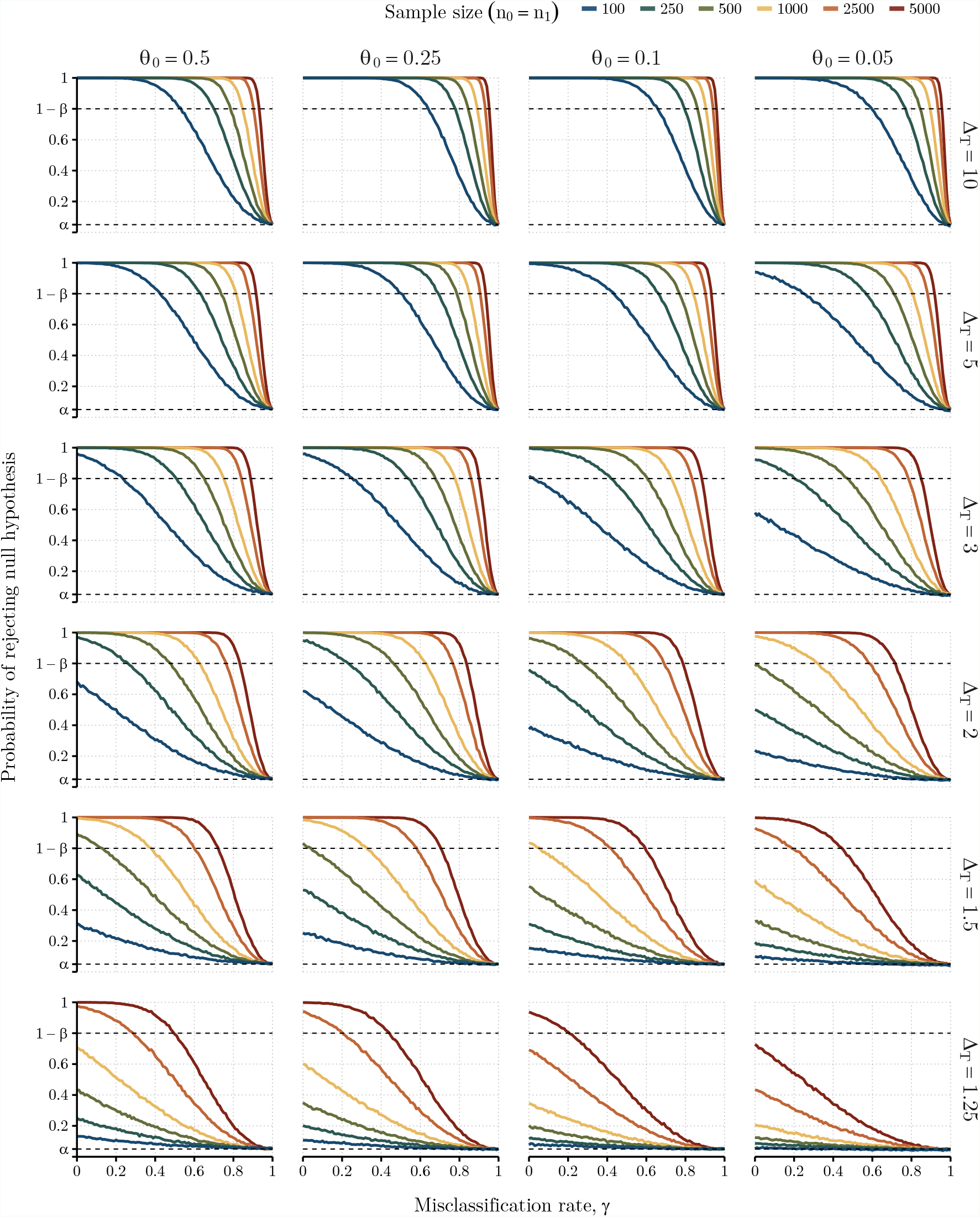
Probabilities of rejecting the null hypothesis, i.e., absence of association between the two populations, as function of the misclassification rate. Each column represents the values attributed to the risk allele frequency found in matched healthy controls and false positive ME/CFS cases (*θ*_0_ ∈ {0.05, 0.1, 0.25, 0.5}). Each row varies the true odds ratio for the association between risk allele frequency assessed between true positive cases and healthy controls (Δ_*T*_ ∈ {1.25, 1.5, 2, 3, 5, 10 }). Power analysis was estimated for different sample sizes of 100, 250, 500, 1000, 2500, and 5000 (*n*_0_ = *n*_1_), represented by the different coloured lines on each scenario. Upper dashed line indicates the target power, where the probability of rejecting the null hypothesis is 1 − *β* = 0.80. Lower dashed line indicates the significance level used, *α* = 0.05.

**Figure 2.**
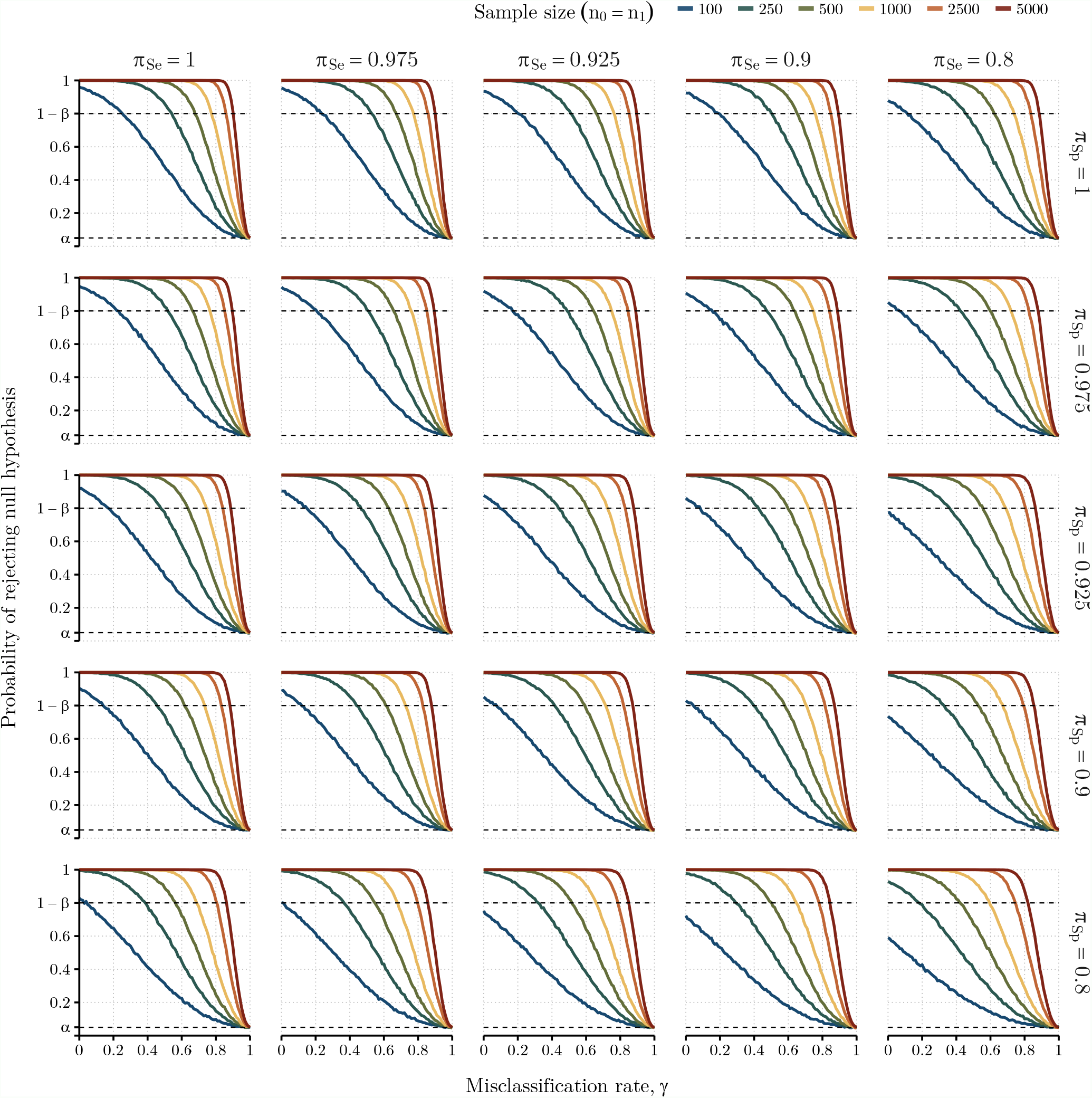
Probabilities of rejecting *H*_0_ as function of the misclassification rate. Each scenario represents simulated results with combination of serology test’s sensitivity, *π*_*se*_, and specificity, *π*_*sp*_, for columns and rows, respectively. Power analysis was estimated for different sample sizes of 100, 250, 500, 1000, 2500, and 5000 (*n*_0_ = *n*_1_), represented by the different coloured lines on each scenario, with probability of exposure in healthy controls fixed as *θ*_0_ = 0.25 and true odds ratio Δ_*T*_ = 3. Upper dashed line indicates the target power, where the probability of rejecting the null hypothesis is 1 − *β* = 0.80. Lower dashed line indicates the significance level used, *α* = 0.05.

As expected, the most optimistic scenarios were associated with Δ_*T*_ = 5 or 10 (i.e., strong associations between the candidate causal factor and true ME/CFS cases). In these scenarios, it was possible to find a maximum misdiagnosis probability in which the target power of 80% was achieved irrespective of the sample size and the probability of the causal factor being present in healthy controls (Table 3). For Δ_*T*_ = 10, a misdiagnosis probability of 0.53 was sufficient to ensure the desired power for sample sizes greater of equal to 100 individuals per study group (*n*_*i*_ ≥ 100), irrespective of *θ*_0_. This minimum probability reduced to 0.24 for Δ_*T*_ = 5. That is, when the association between the candidate causal factor and true ME/CFS cases was strong, the chance of missing its diagnosis could only occur if the chance of ME/CFS misdiagnosis was simultaneously high. Therefore, if the “common disease, rare variant” hypothesis holds true in ME/CFS, it is very likely to be detected by genome-wide association studies.

**Table 3.**
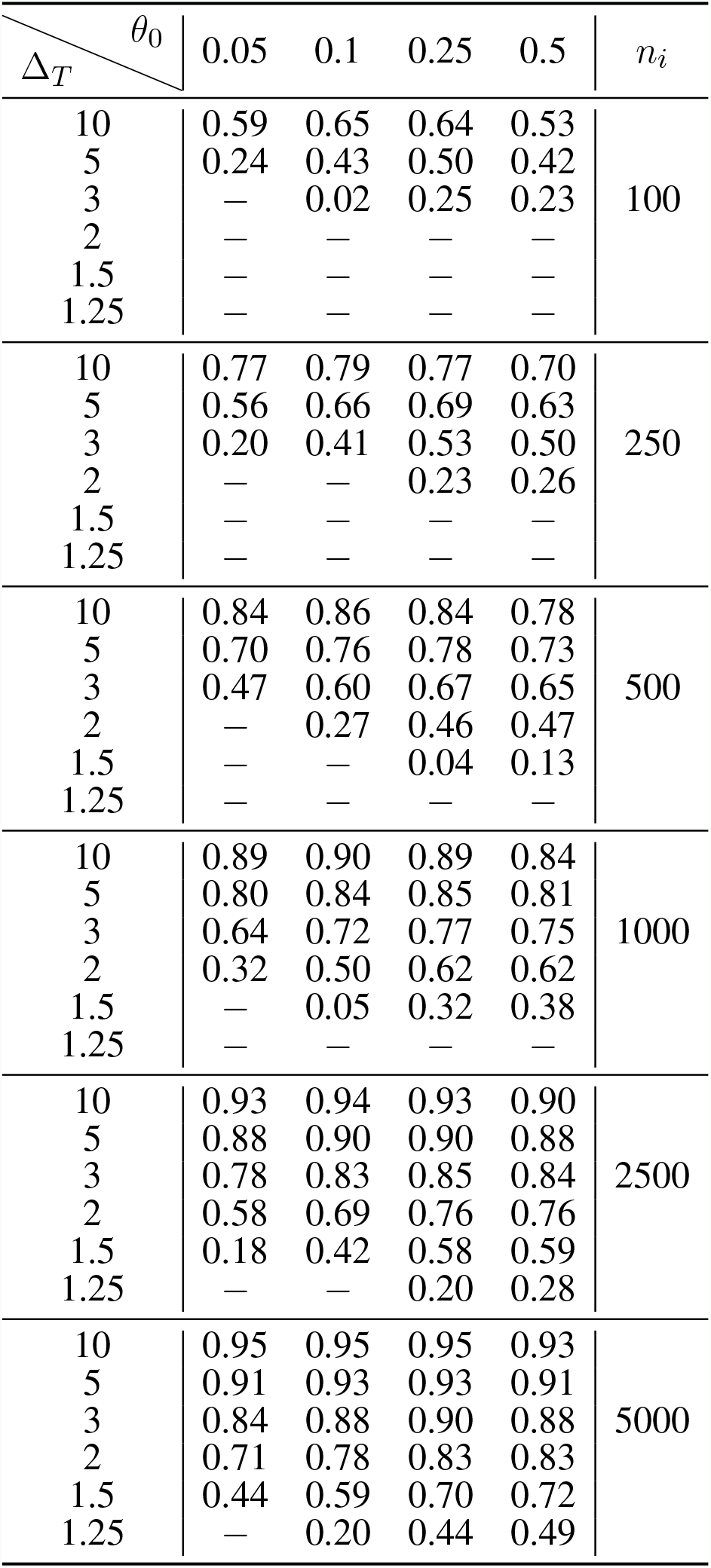
Maximum values of misdiagnosis probability *γ* that still ensured the power of at least 80% to reject the null hypothesis of lack of association at different values of true odds ratio, Δ_*T*_, risk factor probability on healthy controls and false positive cases, *θ*_0_, and sample sizes, *n*_*i*_, *i* = (0, 1). Cells with no value indicate that the desired power could not be reached using the respective parameter combination, even when considering a perfect diagnostic scenario (*γ* = 0).

| $\Delta_T \backslash \theta_0$ | 0.05 | 0.1 | 0.25 | 0.5 | $n_i$ |
| --- | --- | --- | --- | --- | --- |
| 10 | 0.59 | 0.65 | 0.64 | 0.53 | 100 |
| 5 | 0.24 | 0.43 | 0.50 | 0.42 |  |
| 3 | — | 0.02 | 0.25 | 0.23 |  |
| 2 | — | — | — | — |  |
| 1.5 | — | — | — | — |  |
| 1.25 | — | — | — | — |  |
| 10 | 0.77 | 0.79 | 0.77 | 0.70 | 250 |
| 5 | 0.56 | 0.66 | 0.69 | 0.63 |  |
| 3 | 0.20 | 0.41 | 0.53 | 0.50 |  |
| 2 | — | — | 0.23 | 0.26 |  |
| 1.5 | — | — | — | — |  |
| 1.25 | — | — | — | — |  |
| 10 | 0.84 | 0.86 | 0.84 | 0.78 | 500 |
| 5 | 0.70 | 0.76 | 0.78 | 0.73 |  |
| 3 | 0.47 | 0.60 | 0.67 | 0.65 |  |
| 2 | — | 0.27 | 0.46 | 0.47 |  |
| 1.5 | — | — | 0.04 | 0.13 |  |
| 1.25 | — | — | — | — |  |
| 10 | 0.89 | 0.90 | 0.89 | 0.84 | 1000 |
| 5 | 0.80 | 0.84 | 0.85 | 0.81 |  |
| 3 | 0.64 | 0.72 | 0.77 | 0.75 |  |
| 2 | 0.32 | 0.50 | 0.62 | 0.62 |  |
| 1.5 | — | 0.05 | 0.32 | 0.38 |  |
| 1.25 | — | — | — | — |  |
| 10 | 0.93 | 0.94 | 0.93 | 0.90 | 2500 |
| 5 | 0.88 | 0.90 | 0.90 | 0.88 |  |
| 3 | 0.78 | 0.83 | 0.85 | 0.84 |  |
| 2 | 0.58 | 0.69 | 0.76 | 0.76 |  |
| 1.5 | 0.18 | 0.42 | 0.58 | 0.59 |  |
| 1.25 | — | — | 0.20 | 0.28 |  |
| 10 | 0.95 | 0.95 | 0.95 | 0.93 | 5000 |
| 5 | 0.91 | 0.93 | 0.93 | 0.91 |  |
| 3 | 0.84 | 0.88 | 0.90 | 0.88 |  |
| 2 | 0.71 | 0.78 | 0.83 | 0.83 |  |
| 1.5 | 0.44 | 0.59 | 0.70 | 0.72 |  |
| 1.25 | — | 0.20 | 0.44 | 0.49 |  |

Similar optimistic scenarios were observed for sample sizes of 2500 and 5000 individuals per study group with the exception of the case of lowest Δ_*T*_ = 1.25. Combining these large sample sizes with strong associations between the candidate causal factor and the true ME/CFS cases, the target power could only not be achieved when almost all the cases were misdiagnosed (with misdiagnosis probability greater than or equal to 0.88).

The most pessimist scenarios were associated with either Δ_*T*_ = 1.25, 1.5 or *n*_0_ = *n*_1_ = 100. When Δ_*T*_ = 1.25, the sample size had to increase to 2500 or 5000 individuals per group in order to achieve the desired target power. Therefore, for this weak association, the likelihood of finding reproducible results was very low, even under the assumption of a perfect diagnosis. As a consequence, testing the “common disease, common variant hypothesis” in ME/CFS is likely to fail in future genetic associations. Finally, the case of *n*_0_ = *n*_1_ = 100 was particularly problematic given that it was not possible to find any value misdiagnosis probability in which the desired power could be achieved for Δ_*T*_ ≤ 2 (Figure 1).

### 3.2 Simulation study: impact of ME/CFS misdiagnosis and misclassification on the candidate causal factor

We then simulated data of a hypothetical association study in which there were both imperfect diagnoses and misclassification of the candidate causal factor (Figure 2). As explained earlier in this paper, this scenario is the curse of any serological association study in ME/CFS, given the estimation of seropositivity of each individual could be affected by the sensitivity and specificity associated with the classification rule used. At this point, it was clear that for values of Δ_*T*_ = 1.25, 1.5, and 2, the desired power was not often achieved for sample sizes smaller than 500 individuals per group in the case of perfect classification of the causal factor. Therefore, the additional assumption of imperfect classification of the candidate causal factor would make previously estimated power even worse. Because of that, we only performed our simulation study on the more optimistic scenario in which Δ_*T*_ = 3 (Table 4).

**Table 4.** Maximum values of misdiagnosis probability *γ* that still ensures a power of rejecting the null hypotheses of at least 80% for Δ_*T*_ = 3 and *θ*_0_ = 0.25, where *π*_*se*_ and *π*_*sp*_ represent sensitivity and specificity associated with the classification of the candidate, respectively. See Table 3 for more information.

| $\pi_{sp} \backslash \pi_{se}$ | 1 | 0.975 | 0.925 | 0.9 | 0.8 | $n$ |
| --- | --- | --- | --- | --- | --- | --- |
| 1 | 0.25 | 0.23 | 0.20 | 0.19 | 0.11 | 100 |
| 0.975 | 0.22 | 0.20 | 0.17 | 0.15 | 0.06 |  |
| 0.925 | 0.17 | 0.14 | 0.09 | 0.08 | — |  |
| 0.9 | 0.13 | 0.11 | 0.07 | 0.04 | — |  |
| 0.8 | 0.03 | — | — | — | — |  |
| 1 | 0.53 | 0.52 | 0.51 | 0.50 | 0.45 | 250 |
| 0.975 | 0.51 | 0.50 | 0.48 | 0.47 | 0.42 |  |
| 0.925 | 0.47 | 0.46 | 0.43 | 0.42 | 0.36 |  |
| 0.9 | 0.45 | 0.43 | 0.41 | 0.39 | 0.32 |  |
| 0.8 | 0.38 | 0.36 | 0.31 | 0.29 | 0.18 |  |
| 1 | 0.67 | 0.67 | 0.66 | 0.65 | 0.62 | 500 |
| 0.975 | 0.66 | 0.65 | 0.64 | 0.63 | 0.59 |  |
| 0.925 | 0.63 | 0.62 | 0.60 | 0.59 | 0.55 |  |
| 0.9 | 0.61 | 0.61 | 0.59 | 0.57 | 0.52 |  |
| 0.8 | 0.56 | 0.54 | 0.51 | 0.50 | 0.42 |  |
| 1 | 0.77 | 0.77 | 0.76 | 0.75 | 0.73 | 1000 |
| 0.975 | 0.76 | 0.75 | 0.74 | 0.74 | 0.72 |  |
| 0.925 | 0.74 | 0.73 | 0.72 | 0.71 | 0.68 |  |
| 0.9 | 0.73 | 0.72 | 0.71 | 0.70 | 0.67 |  |
| 0.8 | 0.68 | 0.67 | 0.65 | 0.64 | 0.59 |  |
| 1 | 0.85 | 0.85 | 0.85 | 0.84 | 0.83 | 2500 |
| 0.975 | 0.85 | 0.85 | 0.84 | 0.84 | 0.82 |  |
| 0.925 | 0.84 | 0.83 | 0.82 | 0.82 | 0.80 |  |
| 0.9 | 0.83 | 0.82 | 0.81 | 0.81 | 0.79 |  |
| 0.8 | 0.80 | 0.79 | 0.78 | 0.78 | 0.74 |  |
| 1 | 0.90 | 0.90 | 0.89 | 0.89 | 0.88 | 5000 |
| 0.975 | 0.89 | 0.89 | 0.89 | 0.88 | 0.87 |  |
| 0.925 | 0.88 | 0.88 | 0.87 | 0.87 | 0.86 |  |
| 0.9 | 0.88 | 0.87 | 0.87 | 0.87 | 0.85 |  |
| 0.8 | 0.86 | 0.85 | 0.84 | 0.84 | 0.81 |  |

### 3.3 Application to data from two ME/CFS studies

We illustrated the problem of misdiagnosis in data from two ME/CFS studies (20, 21). We started with data from a candidate gene association study (20). In this study, some genetic associations were only found significant when comparing healthy controls to ME/CFS patients with an infectious disease trigger onset (Table 5). The estimated allele-related odds ratios varied from 0.84 [95%CI = (0.56, 1.27)] (rs1799724, *TNF*_2_) to 1.63 [95%CI = (1.04, 2.55)] (rs2476601, *PTPN22*). In our re-analysis, we investigated the impact of misdiagnosis if a replication study were conducted in the same population. In line with the original study, no genotyping errors were assumed for the genetic data. The reported odds ratios were assumed to be the true ones for the population and data were simulated with the same number of reported alleles as in the original study. A description of the simulation procedure can be found elsewhere (10). Similarly to the previous sections, we estimated the probability of rejecting *H*_0_ as a function of the misdiagnosis probability.

**Table 5.**
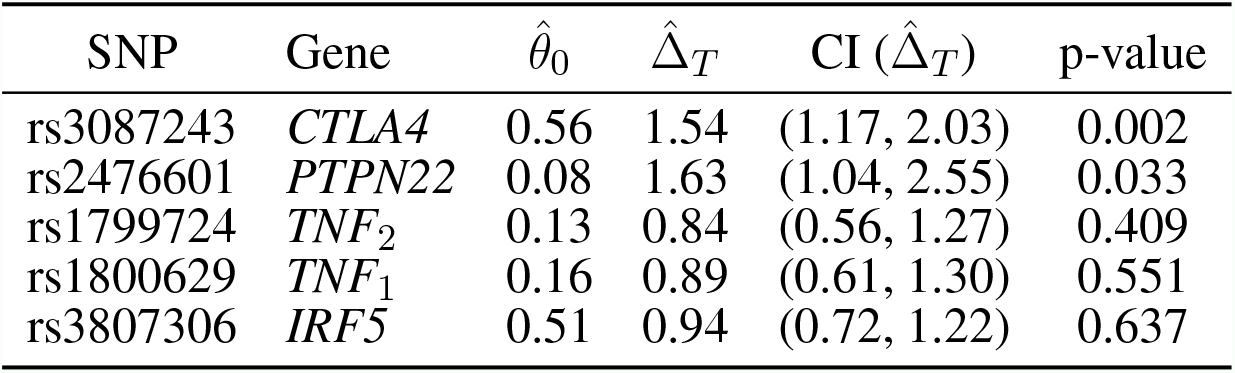
Reported associations of a candidate gene association study (20) where 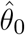 represents the frequencies of the non-reference allele for healthy controls and 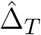is the odds ratio of these allele frequencies when comparing ME/CFS patients with an infectious disease trigger to healthy controls. P-values are associated with the Pearson’s χ^2^ test for 2 × 2 contingency tables.

| SNP | Gene | $\hat{\theta}_0$ | $\hat{\Delta}_T$ | CI ($\hat{\Delta}_T$) | p-value |
| --- | --- | --- | --- | --- | --- |
| rs3087243 | *CTLA4* | 0.56 | 1.54 | (1.17, 2.03) | 0.002 |
| rs2476601 | *PTPN22* | 0.08 | 1.63 | (1.04, 2.55) | 0.033 |
| rs1799724 | *TNF<sub>2</sub>* | 0.13 | 0.84 | (0.56, 1.27) | 0.409 |
| rs1800629 | *TNF<sub>1</sub>* | 0.16 | 0.89 | (0.61, 1.30) | 0.551 |
| rs3807306 | *IRF5* | 0.51 | 0.94 | (0.72, 1.22) | 0.637 |

Again, the estimated probability of rejecting *H*_0_ decreased with the misdiagnosis probability (Figure 3A). More importantly, when the chance of misdiagnosis was low (*γ* < 0.09), it was possible to obtain the target power of 80% for the allele association reported for rs3087243 in *CTLA4*. Therefore, the target power cannot be ensured for *γ >* 0.09. For the remaining SNPs, the target power was never achieved irrespective of the misdiagnosis probability. This is particularly problematic for rs2476601 in *PTPN22* whose association was reported to be significant at a 0.05 significance level. For this SNP, a chance of approximately 10% misdiagnosis deemed a power of about 50%, conceptually making the replication of this result equivalent to flipping a coin.

**Figure 3.**
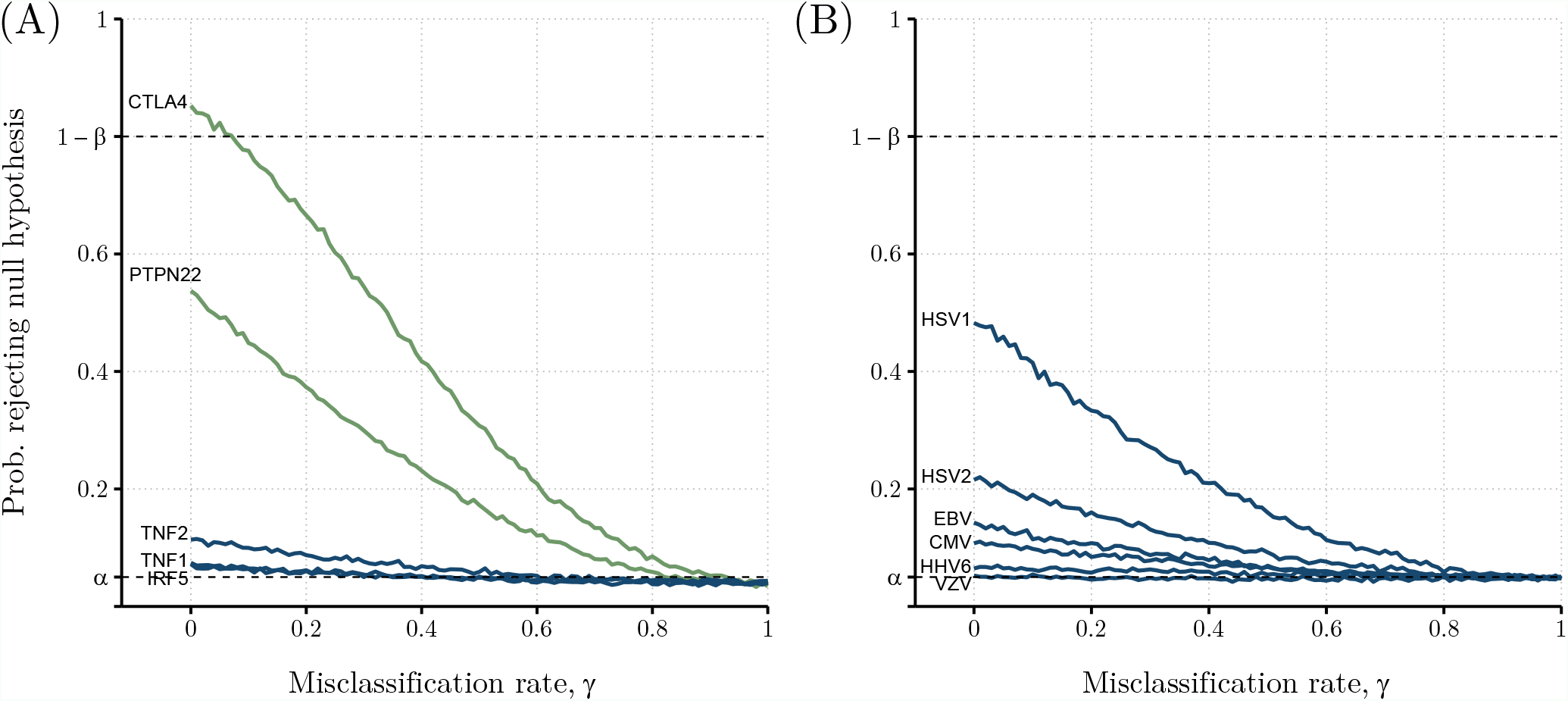
Probabilities of rejecting the *H*_0_ considering real-world data from (A) five different SNPs studied (genes *PTPN22, CTLA4, TNF* (1 and 2), and *IRF5*); and (B) six different human herpes viruses (CMV, EBV, HSV1 and HSV2, VZV, and HHV6), as function of misclassification rate. For each study, risk allele frequencies or probability of exposure and true odds ratio were determined by Steiner et al. (20) (*n*_0_ = 201; *n*_1_ = 305) and Cliff et al. (21) (*n*_0_ = 107; *n*_1_ = 251; *π*_*se*_ = *π*_*sp*_ = 0.975), with determined values shown in Tables 5 and 6, respectively. Green coloured lines indicate candidate risk factors where a significant association with the disease was found in the original study. Blue coloured lines show non-significant ME/CFS risk factors. Upper dashed line indicates the target power, where the probability of rejecting the null hypothesis is 1 − *β* = 0.80. Lower dashed line indicates the significance level used, *α* = 0.05.

The second study referred to putative associations of 6 herpes virus infections with ME/CFS using serological data (21). In these data, all individuals were classified as seronegative or seropositive for each antibody used. Under the assumption of perfect serological classification and diagnosis, the associations of these serological data with severely affected ME/CFS patients ranged from 0.65 [95%CI = (0.21, 1.97)] to 1.60 [95%CI = (0.83, 3.09)] for Epstein-Barr virus (EBV) and Herpes simplex virus 1 (HSV1), respectively (Table 6). For this case, none of the associations was deemed significant at the usual significance level of 5% according to the original study (p-values ≥ 0.16).

**Table 6.**
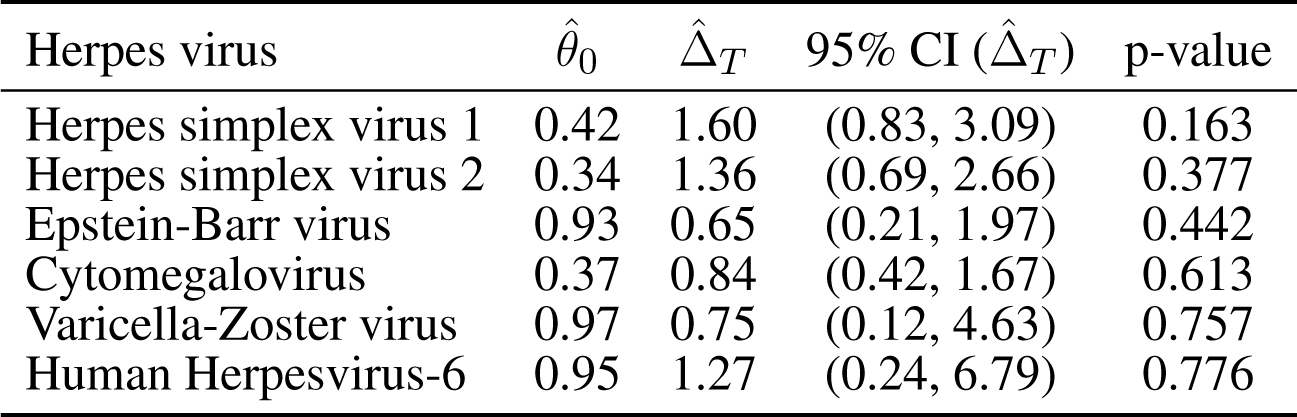
Summary of serological findings from (21), where 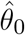 represents the seroprevalence of healthy controls and 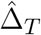*T* refers to the odds ratio for being seropositive when comparing severely-affected ME/CFS patients with healthy controls. The 95% CI 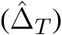 P-values are associated with the Pearson’s χ^2^ test for 2 × 2 contingency tables.

| Herpes virus | $\hat{\theta}_0$ | $\hat{\Delta}_T$ | 95% CI ($\hat{\Delta}_T$) | p-value |
| --- | --- | --- | --- | --- |
| Herpes simplex virus 1 | 0.42 | 1.60 | (0.83, 3.09) | 0.163 |
| Herpes simplex virus 2 | 0.34 | 1.36 | (0.69, 2.66) | 0.377 |
| Epstein-Barr virus | 0.93 | 0.65 | (0.21, 1.97) | 0.442 |
| Cytomegalovirus | 0.37 | 0.84 | (0.42, 1.67) | 0.613 |
| Varicella-Zoster virus | 0.97 | 0.75 | (0.12, 4.63) | 0.757 |
| Human Herpesvirus-6 | 0.95 | 1.27 | (0.24, 6.79) | 0.776 |

The original serological classification was based on a cut-off in the antibody levels determined by the 2*σ*-rule. In general, this cut-off is defined by the mean plus twice the standard deviation of a known seronegative population. Under the assumption of a normal distribution for the seronegative population, the expected specificity of the serological specificity is approximately 0.975 (24). We assumed this value for *π*_*sp*_. For simplicity, we assumed *π*_*se*_ = *π*_*sp*_. Again, we simulated data from this scenario as the original study and estimated the probability of rejecting *H*_0_ as a function of the misdiagnosis probability.

In this study, it was not possible to reach the desired power of 80% to reject *H*_0_ for any of the antibodies (Figure 3B). The best case was observed for the data of antibodies against HSV1. In this case, the maximum power was approximately 0.50 in the absence of misdiagnosis. This power dropped to 0.30 when *γ* = 0.25.

For the remaining cases, the power was almost exclusively less than 0.20. This could partially be explained by the fact that *θ*_0_ is higher than 0.93 for antibody data related to EBV, HHV6, and VZV. Therefore, the chance of finding a significant association is meagre if the study were to be repeated in the same population and under the same conditions.

## 4 DISCUSSION

This study showed the impact of disease misdiagnosis on the reproducibility of associations from a given ME/CFS study. This impact encompasses non-trivial combinations of sample size, effect size (or strength of association), and misdiagnosis probability, in general, plus specificity/sensitivity in the case of serological association studies. In this regard, our simulation study showed that strong associations with ME/CFS can be detected with reasonable power even when the probability of misdiagnosis is not negligible. However, strong associations are unlikely to be detected in ME/CFS, given the persistent difficulty in finding reproducible biomarkers (26) and the lack of consistently significant results reported by different genetic association studies of the disease (29, 30, 31, 32). In addition, even if these strong associations could be detected, the respective estimates should be taken as an underestimation of the true ones due to the presence of apparent cases. In other words, the assumption of misdiagnosis implies that the association estimates are intrinsically biased and, therefore, should be corrected, as done in the estimation of disease prevalence in the presence of imperfect diagnostic tests (15). In general, the chance of misdiagnosis (or misclassification) brings a situation of overparameterization of the respective statistical analysis, where the misdiagnosis probability cannot be estimated from the data directly. To deal with this situation, there are non-trivial solutions in the statistical literature (33, 34). However, the discussion about these advanced solutions was out of the scope of this study.

It was also clear that sufficiently large sample sizes can compensate for the reduction in power due to misdiagnosis and misclassification of a potential causal factor. In this regard, a minimum sample size of 500 individuals per group minimises the negative impact of misdiagnosis alone on power. This minimum sample size required an increase towards 1000 individuals per group when the misdiagnosis probability is combined with the misclassification probabilities associated with the potential causal factor. In general, the sampling of such large numbers of individuals is challenging, but is becoming feasible in common, well-recognised, and highly-funded diseases, such as cancer, cardiovascular diseases (35), and autoimmune disorders (36, 37). However, for ME/CFS, where the research is affected by limited funding and poor societal recognition (38), the recruitment of large numbers of patients might be problematic. A possible solution to this problem is to use a collection of samples available from existing biobanks. In this regard, the UK ME/CFS Biobank is fully dedicated to the advancement and acceleration of ME/CFS research. It comprises biological samples of more than 500 individuals, including ME/CFS patients, healthy controls, and patients with multiple sclerosis as an additional control group (39). The strength of this biobank is a detailed characterisation of all the participants and the use of current standards for ME/CFS diagnosis. However, this biobank only contemplates around 250 ME/CFS patients. As an alternative, the UK Biobank has been reported to include 1890 to 2105 ME/CFS cases (30, 32). However, it is unclear the accuracy of the respective data given that ME/CFS diagnosis was self-reported by participants. In this scenario, the most obvious solution to increase the sample size is to conduct multi-centric studies, as suggested by the European ME/CFS Biomarker Landscape project (26). Such a research strategy has already been adopted by a recent genetic association study that contemplated Norwegian, Danish, and British cohorts at different stages of the analysis (32). Notwithstanding their attractiveness and potential impact on biomarker discovery, multi-centric studies require considerable funding, robust research protocols and ethics, and strong compliance among the participating sites. In this scenario, the major challenge of these studies consists in the fact that ME/CFS research is currently underfunded (40, 41). However, there is hope emerging from initiatives such as EUROMENE and the US Clinician Coalition, whose objective is to promote and foster collaboration and shared resources among their members. In addition, the similarity between long-Covid and ME/CFS highlighted the need to increase research funding on these conditions to mitigate their economic costs in the future (42). Therefore, large multi-centric studies in ME/CFS are expected to become a routine research practice in the future.

The potential misclassification of a binary causal factor is often neglected in the analysis of both genetic and serological association studies of ME/CFS. In previous studies, this neglect does not seem problematic because estimated genotype error rates are below 1% (43). However, higher genotype error rates can be obtained for rare genetic markers (44), and genetic markers with minor allele frequencies less than 0.01 or 0.05 are typically excluded from genetic association studies of ME/CFS (22, 32, 45). The parameterization of our simulation study (i.e., *θ*_0_ *>* 0.05) is then in line with this research practice and, thus, the respective findings are directly applicable for understanding the reproducibility of current genetic association studies of ME/CFS. Within this scenario, Dibble et al. (31) advocated for conducting well-powered studies with large cohort sizes. In the present context, such a suggestion is not only important to detect common variants with low genetic effects, but also to minimise the impact of misdiagnosis on the underlying power function, as demonstrated by the present study. In the case of serological association studies, the serological status of each individual can only be determined statistically. Generally, when the antibody distributions of the underlying seronegative or seropositive populations do not overlap with each other, the serological status of each individual can be determined almost perfectly. In practice, serological data of herpesviruses from the UK ME/CFS Biobank showed that the distributions of these serological populations can overlap with each other, as demonstrated by data related to herpesviruses serology from the UK ME/CFS Biobank (19). However, current serological association studies do not report the sensitivity and specificity of the corresponding serological classification rule and, therefore, it is unclear how these misclassification parameters vary from one study to another and how they affect the power of detecting underlying associations. The situation is problematic given that these studies are also affected by the high seroprevalence of many viruses in the population, the possibility of batch and laboratory effects, differences in the antigens used, interactions between serological outcomes, and typical confounders, such as gender and age, and the use of different criteria for seropositivity estimation (19, 46, 47). Therefore, it is imperative the reporting of the uncertainty associated with the serological classification rule in future studies.

In this study, we assumed independence between disease association and possible confounding factors. In practice, this assumption might not always be true, given the evidence that age, gender, or exposure to a given infectious agent can influence the respective findings (19, 48). Hence, the above assumption is more appropriate for interpreting studies in which individuals are sampled from a specific combination of confounding factors. Such a sampling strategy is particularly adopted in DNA methylation studies in which adult women with matched age and BMI are the target population (49).

Furthermore, we assumed no misdiagnosis of healthy controls. This assumption deserves a comment in light of current data and research practices. On the one hand, ME/CFS patients and some healthy controls share the same symptoms and level of fatigue (10, 50). On the other hand, studies such as Loebel et al. 17 and Szklarski et al. (48) recruited healthy individuals solely based on self-reporting. Such recruiting strategy, although reducing research costs, increases the chance of sampling asymptomatic but unhealthy individuals. A similar problem can be expected in Kaushik (51), Johnston et al. (52), and Lande et al. (53), using control samples from blood donors. In Herrera et al. (29), Blauensteiner et al. (54), de Vega et al. (55), and Trivedi et al. (56), both asymptomatic non-obese and obese individuals were recruited for subsequent matching with ME/CFS patients. However, some of these individuals can be seen as cases of undiagnosed metabolic syndrome (57, 58, 59). Recruiting these individuals is particularly problematic given the association of metabolic syndrome with ME/CFS (60). With all of the evidence, healthy controls could also be divided into true and apparent controls, thus further reducing the statistical power to detect a putative disease association. Therefore, the most general assumption is to consider the possibility in which undiagnosed patients of ME/CFS or any other disease can be mistakenly recruited as healthy controls. To avoid this situation, the health status of putative controls should also be routinely confirmed with lab testing.

We formulated the statistical problem in terms of ME/CFS misdiagnosis, considering the suspected ME/CFS cohort to be composed of misdiagnosed and true patients. From a modelling standpoint, our formulation can be reframed as a related problem in which the ME/CFS group can be divided into two subgroups of patients, and the association to be detected is only present in one of these subgroups. This alternative formulation follows the idea that ME/CFS encompasses multiple disease subtypes (61). However, we should be cautious when interpreting our results in this alternative scenario. First, the assumption of only two disease subgroups seems overly simplistic, given that it was suggested more than seven genomic subtypes for the disease (62, 63). Second, these potential subtypes might overlap with each other, given that no clear stratification of patients could be identified using symptoms’ data (10). Hence, the membership of each patient to a given disease subtype might be probabilistic in nature. However, this situation is more complex and was not captured by the presented mathematical formulation.

In summary, this study discusses the reproducibility of ME/CFS studies in terms of disease misdiagnosis. Given the absence of an objective biomarker for disease diagnosis, the most obvious recommendations are: (i) to use recommended case definitions for ME/CFS diagnosis; (ii) to increase cohort sizes using multi-centric studies; and (iii) to confirm the health status of putative controls. These recommendations combined are likely to minimise the impact of misdiagnosis of both patients and controls on the disease associations to be detected. A final recommendation is to perform post-hoc power calculations as a function of misdiagnosis probability in order to quantify the likelihood of a reproducible finding in the presence of imperfect diagnosis of ME/CFS.

## Data Availability

The source code and datasets generated can be found in the GitHub public repository (https://github.com/jtmalato/misclassification-simulations).

## AUTHOR CONTRIBUTIONS

JM and NS conceived and designed the analysis. JM performed the computer simulations, statistical analysis, and wrote the initial manuscripts. All authors participated actively in the discussion of the results. All authors have read, revised, and approved the final version of the manuscript.

## ACKNOWLEDGMENTS

JM would like to thank Prof. Przemysław Biecek and the MI^2^ DataLab group at Faculty of Mathematics and Information Science, Warsaw University of Technology, Warsaw, Poland, for their support during part of the analysis and discussion of this article.

## DATA AVAILABILITY STATEMENT

All simulations and analyses were done using R statistical software, version 4.1.0 (64). The source code and datasets generated can be found in the GitHub public repository (https://github.com/jtmalato/misclassification-simulations).

